# Human versus Artificial Intelligence: ChatGPT-4 Outperforming Bing, Bard, ChatGPT-3.5, and Humans in Clinical Chemistry Multiple-Choice Questions

**DOI:** 10.1101/2024.01.08.24300995

**Authors:** Malik Sallam, Khaled Al-Salahat, Huda Eid, Jan Egger, Behrus Puladi

**Author notes:** **Corresponding Author:** (MS).

## Abstract

The advances in large language models (LLMs) are evolving rapidly. Artificial intelligence (AI) chatbots based on LLMs excel in language understanding and generation, with potential utility to transform healthcare education and practice. However, it is important to assess the performance of such AI models in various topics to highlight its strengths and possible limitations. Therefore, this study aimed to evaluate the performance of ChatGPT (GPT-3.5 and GPT-4), Bing, and Bard compared to human students at a postgraduate master’s (MSc) level in Medical Laboratory Sciences. The study design was based on the METRICS checklist for the design and reporting of AI-based studies in healthcare. The study utilized a dataset of 60 Clinical Chemistry multiple-choice questions (MCQs) initially conceived for assessment of 20 MSc students. The revised Bloom’s taxonomy was used as the framework for classifying the MCQs into four cognitive categories: Remember, Understand, Analyze, and Apply. A modified version of the CLEAR tool was used for assessment of the quality of AI-generated content, with Cohen’s κ for inter-rater agreement. Compared to the mean students’ score which was 40/60 (66.8%), GPT-4 scored 54/60 (90.0%), followed by Bing (46/60, 76.7%), GPT-3.5 (44/60, 73.3%), and Bard (40/60, 66.7%). Statistically significant better performance was noted in lower cognitive domains (Remember and Understand) in GPT-3.5, GPT-4, and Bard. The CLEAR scores indicated that ChatGPT-4 performance was “Excellent” compared to “Above average” performance of ChatGPT-3.5, Bing, and Bard. The findings indicated that ChatGPT-4 excelled in the Clinical Chemistry exam, while ChatGPT-3.5, Bing, and Bard were above-average. Given that the MCQs were directed to postgraduate students with a high degree of specialization, the performance of these AI chatbots was remarkable. Due to the risks of academic dishonesty and possible dependence on these AI models, the appropriateness of MCQs as an assessment tool in higher education should be re-evaluated.

## Introduction

The domain of higher education is set for a new transformative era [1, 2]. This transformation will be driven by the infiltration of artificial intelligence (AI) into various academic aspects [3–7]. Specifically, the incorporation of AI into higher education can help in enhancing personalized learning, supporting research, automating the grading, facilitating the human-computer interaction, time-saving assistance, and enhancing the students’ satisfaction [8–12].

Nevertheless, the AI utility in higher education does not only hold promising opportunities but also valid concerns, both of which warrant critical and robust examination [13–15]. This research endeavor is necessary to guide the ethical, responsible, and productive use of AI to enhance higher education guided by a robust scientific evidence [14, 16, 17]. The relevance of the quest to meticulously examine the benefits and challenges of AI in higher education is also important in light of the current evidence showing that a substantial number of university students are already using AI chatbots [18–22].

Despite the benefits of AI in higher education, it simultaneously raises valid concerns about the academic integrity [23, 24]. The ease with which AI can perform complex tasks might inadvertently encourage academic dishonesty, potentially undermining the educational ethics [23, 25]. Furthermore, the reliance on AI for academic tasks could trigger a decline in critical thinking and personal development skills among students, both of which are essential outcomes to enable the graduates in achieving economic, technological, and social advancements [26, 27].

Ultimately, with notable capabilities of AI chatbots in understanding and engaging in helpful conversations, would usher a paradigm shift in higher education [8, 14, 28]. This AI-driven change could be a key moment in educational history, with impact surpassing the advent of the internet and the transition to online teaching [16, 17, 29]. Therefore, the stakeholders in the academia must strike the right balance between embracing technological innovations while preserving the core values of education [30–32]. Thus, the integration of AI into higher education is inevitable, and the academic organizations must adapt to this evolution [3]. This adaptation involves the need to emphasize educational aspects such as self-reflection, critical thinking, problem-solving, and independent learning [33]. Consequently, the educational systems can benefit from AI as a tool to complement, rather than replace, human intellect and creativity [34].

In the quest of transition to the AI era in education, guidance by robust scientific evidence is crucial. One of the primary steps in this process is to scientifically evaluate the performance of the commonly used and popular AI tools, such as ChatGPT (by OpenAI, San Francisco, CA), Bing (by Microsoft Corporation, Redmond, WA), and Bard (by Google, Mountain View, CA). Several recent studies explored the performance of AI-based models in multiple-choice questions (MCQs), particularly within a broad spectrum of healthcare fields as recently reviewed by Newton and Xiromeriti [35]. The observed variability in AI performance can be ascribed to several factors, including the different AI models tested, varying approaches to prompting, language variations, and the diversity of the topics tested, among others [35–37]. Thus, continued investigation into this research area is needed to elucidate the determinants of AI model performance across various dimensions which can guide improvements in AI algorithms. However, it is essential that such explorations are conducted utilizing a standardized, refined methodology [36, 38].

The use of multiple-choice questions (MCQs) have traditionally been fundamental as an objective approach in academic evaluation [39]. The versatility of MCQs is shown through the use of the Bloom’s taxonomy and its subsequent revised framework [40, 41]. The Bloom’s taxonomy can guide structuring MCQs to align with specific cognitive functions needed to provide correct answers [42]. This alignment is key in assessing the students’ achievement of the intended learning outcomes [43]. The taxonomy stratifies these cognitive functions into distinct categories. The lower cognitive levels encompass knowledge, which emphasizes “Recall”, and comprehension, centered on “Understanding”. Conversely, the higher cognitive functions include “Apply”, key in problem-solving, and “Analyze”, entailing the systematic breakdown of information [40, 41].

In the context of assessing the performance of AI model performance in MCQs based on the Bloom’s taxonomy, a pioneering study by Herrmann-Werner et al. assessed ChatGPT-4 with 307 psychosomatic medicine MCQs [44]. The study demonstrated ChatGPT-4 ability to pass the exam irrespective of the prompting method. Notably, cognitive errors were more prevalent in “Remember” and “Understand” categories. Another recent study demonstrated that ChatGPT-3.5 correctly answered 64 of 80 medical microbiology MCQs, albeit below student averages, with better performance in the “Remember” and “Understand” categories and more frequent errors in MCQ with longer choices in terms of word count [45].

In this study, the objective was to synthesize and expand upon recent research examining the performance of AI chatbots in various examinations. This research was informed by seminal studies, such as the Kung et al. evaluation of ChatGPT in the United States Medical Licensing Examination (USMLE) [37], and also it aimed to extend the evidence of AI chatbot performance in a topic rarely encountered in literature, namely the Clinical Chemistry at postgraduate level. The novel contribution of the current study lies in employing a standardized framework, termed “METRICS” for the design and reporting of AI assessment studies, coupled with an in-depth analysis of AI models’ rationale behind responses, using an evaluation tool specifically tailored for AI content evaluation referred to as “CLEAR” [36, 38].

The study hypothesized that postgraduate students, particularly in the field of clinical chemistry, will demonstrate superior performance compared to AI models. We anticipate that this disparity will be especially evident in tasks requiring higher cognitive functions, such as “Apply” and “Analyze”. This study aimed to critically assess the current capabilities of AI in an academic setting and explore the differences of human versus artificial intelligence in complex problem-solving scenarios.

## Materials and Methods

### Study design

The study utilized the METRICS checklist for the design and reporting of AI studies in healthcare [36]. The basis of the study was a dataset of 60 MCQs, used in a Clinical Chemistry examination. This examination was part of the Medical Laboratory Sciences Clinical Chemistry course, tailored for Master of Science (MSc) students in Medical Laboratory Sciences at the School of Medicine, University of Jordan.

The specific exam in focus was conducted in-person and 20 students undertook the examination during the Autumn Semester of the 2019/2002 academic year. The students’ performance in each question was available for comparison with AI models.

The MCQs utilized in this exam were designed by the first author (M.S.), who is a Jordan Medical Council (JMC) certified consultant in Clinical Pathology. Additionally, the first author (M.S.) has been a dedicated instructor for this course since the Academic Year 2018/2019. The MCQs were original, ensuring there were no copyright concerns.

### Ethical considerations

In conducting this study, we paid careful attention to ethical implications. Ethical clearance for this research was determined to be non-essential, given the nature of the data involved. The data utilized were entirely anonymized, ensuring no breach of confidentiality or personal privacy. Additionally, the university examination results, which formed part of our dataset, are publicly accessible and open for academic scrutiny. Moreover, the MCQs employed in the study were originally created by the first author. These questions are devoid of any copyright concerns, further reinforcing the ethical integrity of our research approach.

### MCQ features and indices of human students’ performance

The indices of student performance included facility index defined as the proportion of students who correctly answered the MCQ divided by the total number of students (*n*=20). The students were then divided into the upper group comprising the top 5 performing students, and the middle group comprising the middle 10 students and the lower group comprising the lower scoring 5 students. The “Discrimination Index” (DI) was then calculated based on the difference between the percent of correct responses in the upper group and the percent of correct responses in the lower group. This was followed by the calculation of the “Maximum Discrimination” based on the sum of the percent in the upper and lower groups marking the item correctly. Then, the Discrimination Efficiency (DE) of the MCQ was calculated as the ratio of DI to the Maximum Discrimination. The classification of the MCQs based on the revised Bloom’s taxonomy four cognitive levels “Remember”, “Understand”, “Apply”, and “Analyze” was based on a consensus between the first and second authors, both of which are certified Clinical Pathologists.

### Models of AI tested, settings, testing time, and duration

In this study, a detailed evaluation of four AI models was conducted, each selected for its relevance, popularity, and advanced capabilities in language processing as follows: First, ChatGPT-3.5 (OpenAI, San Francisco, CA) [46]: This model is grounded in the GPT-3.5 architecture deployed using its default settings and was assessed as of its latest update at time of testing as of January 2022.

Second, ChatGPT-4 (OpenAI, San Francisco, CA) [46]: An advancement in the Generative Pre-trained Transformer (GPT) series, with the most recent update from April 2023 at time of testing. Third, Bing Chat (GPT-4 Turbo) [47]: This model uses the GPT-4 Turbo model. At the time of testing, the version was updated until April 2023 and we selected the more balanced conversation style. Fourth, Bard (Google, Mountain View, CA) [48]: This Google AI GPT model was last updated on October 4, 2023, at time of testing.

The testing of these models was conducted over a concise period, spanning November 27 to November 28, 2023. Our methodological approach involved initiating interactions with GPT-3.5, GPT-4, and Bard using a single page. For Bing Chat, we used the “New Topic” option considering the limit of responses posed by this model (50 at maximum). Additionally, we opted not to use the “regenerate response” feature in ChatGPT and abstained from providing feedback in all models to avoid feedback bias.

### Prompt and language Specificity

In this study, we meticulously crafted the prompts used for interacting with the AI models to ensure clarity and consistency in the testing process. For ChatGPT-3.5, ChatGPT-4, and Bard, the following exact prompt was used: “For the following 60 Clinical Chemistry MCQs that will be provided one by one, please select the most appropriate answer for each MCQ, with an explanation for the rationale behind selecting this choice and excluding the other choices. Please note that only one choice is correct while the other four choices are incorrect. Please note that these questions were designed for masters students in medical laboratory sciences.” This was followed by prompting each MCQ one by one. For Bing, the following prompt was used for each MCQ: “For the following 60 Clinical Chemistry MCQs that will be provided one by one, please select the most appropriate answer for each MCQ, with an explanation for the rationale behind selecting this choice and excluding the other choices. Please note that only one choice is correct while the other four choices are incorrect. Please note that these questions were designed for masters students in medical laboratory sciences.”

All MCQs were presented in English. This choice was based on the fact that English is the official language of instruction for the MSc program in Medical Laboratory Sciences at the University of Jordan.

### AI content evaluation approach and individual involvement in evaluation

First, we objectively assessed the correctness of responses based on the key answers of the MCQs. Then, subjective evaluation of the AI generated content was based on a modified version of the CLEAR tool. This involved assessing the content on three dimensions as follows: First, completeness of the generated response. Second, accuracy reflected by lack of false knowledge and the content being evidence-based. Third, appropriateness and relevance of content being easy to understand, well organized, and free from irrelevant content [38]. Each dimension was scored on a 5-point Likert scale ranging from 1=poor, 2=satisfactory, 3=good, 4=very good, to 5=excellent. A list of the key points to be considered in the assessment was set beforehand to increase objectivity.

The content generated by the four models was evaluated by two raters independently; the first author (M.S.) a consultant in Clinical Pathology, and the second author (K.A.) a specialist in Clinical Pathology, both certified in Clinical Pathology from the Jordan Medical Council (JMC).

### Data source transparency and topic range

The MCQs were totally conceived by the first author and sole instructor of the course. Sources of the material taught during the course were the following three textbooks: Tietz Textbook of Clinical Chemistry and Molecular Diagnostics; Clinical Chemistry: Principles, Techniques, and Correlations; and Henry’s Clinical Diagnosis and Management by Laboratory Methods [49–51].

The scope of topics covered in the MCQs were as follows: Adrenal Function, Amino Acids and Proteins, Body Fluid Analysis, Clinical Enzymology, Electrolytes, Gastrointestinal Function, Gonadal Function, Liver Function, Nutrition Assessment, Pancreatic Function, Pituitary Function, Thyroid Gland, and Trace Elements.

### Statistical and data analyses

The statistical analysis was conducted using IBM SPSS Statistics Version 26.0 (Armonk, NY: IBM Corp). The continuous variables were presented as means and standard deviations (SD), while categorical data were summarized as frequencies and percentages [N (%)]. To explore the associations between categorical variables, we employed the chi-squared test (χ^2^), while to explore the associations between scale variables and categorical variables, non-parametric tests were utilized: the Mann–Whitney *U* test (M-W) and the Kruskal Wallis test (K-W). The Kolmogorov-Smirnov test was employed to confirm the non-normality of the scale variables: facility index (FI, *P*=.042), discriminative efficiency (DE, *P*=.011), word count for both stem and choices (*P*<.001 for both), average completeness, accuracy/evidence, appropriateness/relevance, and the mCLEAR scores (*P*<.001 for the four scores). *P* values <0.050 were considered statistically significant. For multiple comparisons, post hoc analysis was conducted using the M-W test. To account type I error due to multiple comparisons, we adjusted the α level using the Bonferroni correction. Consequently, the adjusted α level for conducting pairwise comparisons between the four AI models was set at *P*=0.0083.

The MCQs were categorized based on the FI as “difficult” for an FI of 0.40 or less, “average” for an FI > 0.40 and ≤ 0.80, and “easy” for an FI > 0.80. Additionally, the DE was stratified into “poor discrimination” if the DE was between −1 to zero, “satisfactory discrimination” for DE > zero to < 0.40 as satisfactory, and DE ≥ 0.4 indicating “good discrimination”.

The inter-rater agreement was assessed using Cohen’s kappa (κ) values, which ranged from very good to excellent. For ChatGPT-3.5, the agreement was κ=0.874 for Completeness, κ=0.921 for Accuracy, and κ=0.723 for Relevance. ChatGPT-4 showed κ=0.845 for Completeness, a perfect κ=1 for Accuracy, and κ=0.731 for Relevance. Bing displayed κ values of 0.911 for Completeness, 0.871 for Accuracy, and 0.840 for Relevance. Lastly, Bard’s agreement was κ=0.903 for Completeness, κ=1 for Accuracy, and κ=0.693 for Relevance. Finally, the overall modified CLEAR (mCLEAR) scores for AI content quality were averaged based on the scores of the two raters and categorized as: “Poor” (1–1.79), “Below average” (1.80–2.59), “Average” (2.60–3.39), “Above average” (3.40–4.19), and “Excellent” (4.20– 5.00) similar to the previous approach in [52].

## Results

### Overall performance of the tested AI models compared to the human students

The overall performance of the MSc students in the exam was reflected in the average score of 40.05±7.23 (66.75%), with the range of scores of the students of 24–54 (40.00%–90.00%). The performance of the four AI models varied with the best performance for ChatGPT-4 scoring 54/60 (90.00%), followed by Bing scoring 46/60 (76.67%), ChatGPT-3.5 scoring 44/60 (73.33%), and finally Bard scoring 40/60 (66.67%).

### Human students’ performance based on the revised Bloom’s taxonomy

The MCQ metrics were derived from the performance of the 20 MSc students in the exam. The best performance was in the “Remember” category, followed by the “Apply” category, “Understand” category, while the worst performance was in the “Analyze” category; however, these differences lacked statistical significance (**Table 1**).

**Table 1.** Multiple-choice questions (MCQs) metrics stratified by the revised Bloom’s taxonomy as derived from the performance of 20 MSc students.

| **Revised Bloom's taxonomy** | **Remember** | **Understand** | **Apply** | **Analyze** | **P value<sup>3</sup>** |
| --- | --- | --- | --- | --- | --- |
| MCQ metric | Mean±SD<sup>2</sup> | Mean±SD | Mean±SD | Mean±SD |  |
| Facility index | 0.74±0.22 | 0.61±0.28 | 0.71±0.21 | 0.6±0.17 | .180 |
| Discriminative efficiency | 0.24±0.25 | 0.24±0.27 | 0.17±0.43 | 0.43±0.41 | .482 |
| MCQ stem word count | 15.04±6.5 | 24±16.95 | 73.4±57.06 | 25.31±27.55 | .052 |
| MCQ choices word count | 13.5±9.29 | 24.83±17.76 | 22.2±8.07 | 29.54±28.64 | .153 |
| **Revised Bloom's cognitive level<sup>1</sup>** | **Lower** |  | **Higher** |  | **P value<sup>4</sup>** |
| Facility index | 0.68±0.25 |  | 0.63±0.18 |  | .225 |
| Discriminative efficiency | 0.24±0.25 |  | 0.36±0.42 |  | .205 |
| MCQ stem word count | 18.88±12.77 |  | 38.67±42.34 |  | .265 |
| MCQ choices word count | 18.36±14.54 |  | 27.5±24.61 |  | .268 |

<sup>1</sup>Lower cognitive level includes “Remember” and “Understand” categories, while the higher cognitive level includes “Apply” and “Analyze” categories; <sup>2</sup>SD: Standard deviation; <sup>3</sup>Calculated using the Kruskal Wallis test; <sup>4</sup>Calculated using the Mann Whiteny *U* test.

### Performance of the AI models based on the MCQ metrics

The performance of the four tested AI models was stratified based on the MCQ metrics. Significantly lower number of correct answers was seen in difficult MCQs in both Bing and Bard (**Table 2**), while the MCQ stem and choices word counts were not associated with AI models’ performance.

**Table 2.**
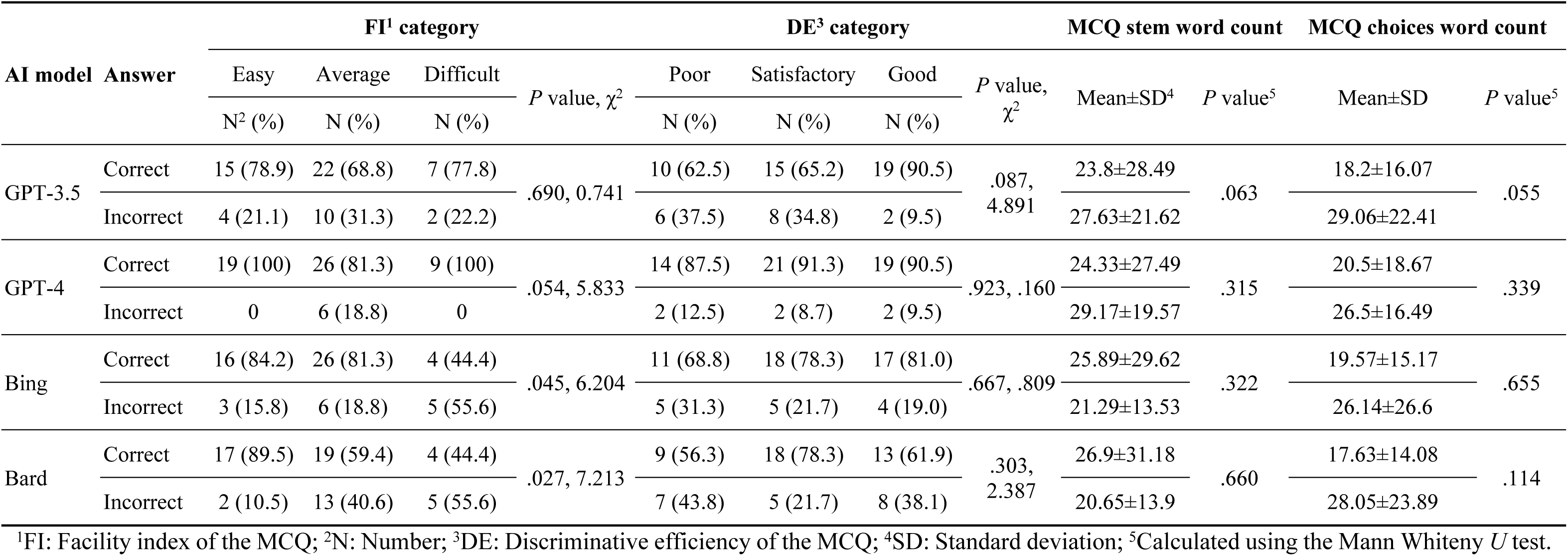
Artificial intelligence (AI)-based model performance based on the multiple-choice question (MCQ) metrics.

| AI model | Answer | FI<sup>1</sup> category |  |  |  | DE<sup>3</sup> category |  |  |  | MCQ stem word count |  | MCQ choices word count |  |
| --- | --- | --- | --- | --- | --- | --- | --- | --- | --- | --- | --- | --- | --- |
| | | Easy | Average | Difficult | *P* value, $\chi^2$ | Poor | Satisfactory | Good | *P* value, $\chi^2$ | Mean±SD<sup>4</sup> | *P* value<sup>5</sup> | Mean±SD | *P* value<sup>5</sup> |
|  |  | N<sup>2</sup> (%) | N (%) | N (%) |  | N (%) | N (%) | N (%) |  |  |  |  |  |
| GPT-3.5 | Correct | 15 (78.9) | 22 (68.8) | 7 (77.8) | .690, 0.741 | 10 (62.5) | 15 (65.2) | 19 (90.5) | .087, 4.891 | 23.8±28.49 | .063 | 18.2±16.07 | .055 |
|  | Incorrect | 4 (21.1) | 10 (31.3) | 2 (22.2) |  | 6 (37.5) | 8 (34.8) | 2 (9.5) |  | 27.63±21.62 |  | 29.06±22.41 |  |
| GPT-4 | Correct | 19 (100) | 26 (81.3) | 9 (100) | .054, 5.833 | 14 (87.5) | 21 (91.3) | 19 (90.5) | .923, .160 | 24.33±27.49 | .315 | 20.5±18.67 | .339 |
|  | Incorrect | 0 | 6 (18.8) | 0 |  | 2 (12.5) | 2 (8.7) | 2 (9.5) |  | 29.17±19.57 |  | 26.5±16.49 |  |
| Bing | Correct | 16 (84.2) | 26 (81.3) | 4 (44.4) | .045, 6.204 | 11 (68.8) | 18 (78.3) | 17 (81.0) | .667, .809 | 25.89±29.62 | .322 | 19.57±15.17 | .655 |
|  | Incorrect | 3 (15.8) | 6 (18.8) | 5 (55.6) |  | 5 (31.3) | 5 (21.7) | 4 (19.0) |  | 21.29±13.53 |  | 26.14±26.6 |  |
| Bard | Correct | 17 (89.5) | 19 (59.4) | 4 (44.4) | .027, 7.213 | 9 (56.3) | 18 (78.3) | 13 (61.9) | .303, 2.387 | 26.9±31.18 | .660 | 17.63±14.08 | .114 |
|  | Incorrect | 2 (10.5) | 13 (40.6) | 5 (55.6) |  | 7 (43.8) | 5 (21.7) | 8 (38.1) |  | 20.65±13.9 |  | 28.05±23.89 |  |

<sup>1</sup>FI: Facility index of the MCQ; <sup>2</sup>N: Number; <sup>3</sup>DE: Discriminative efficiency of the MCQ; <sup>4</sup>SD: Standard deviation; <sup>5</sup>Calculated using the Mann Whiteny *U* test.

### Performance of the AI models based on the revised Bloom’s taxonomy

Upon analyzing the AI models’ performance in MCQs stratified per the four revised Bloom’s categories, only ChatGPT-4 showed statistically significant better performance in the Remember and Understand categories compared to Apply and Analyze categories (Table 3).

**Table 3.**
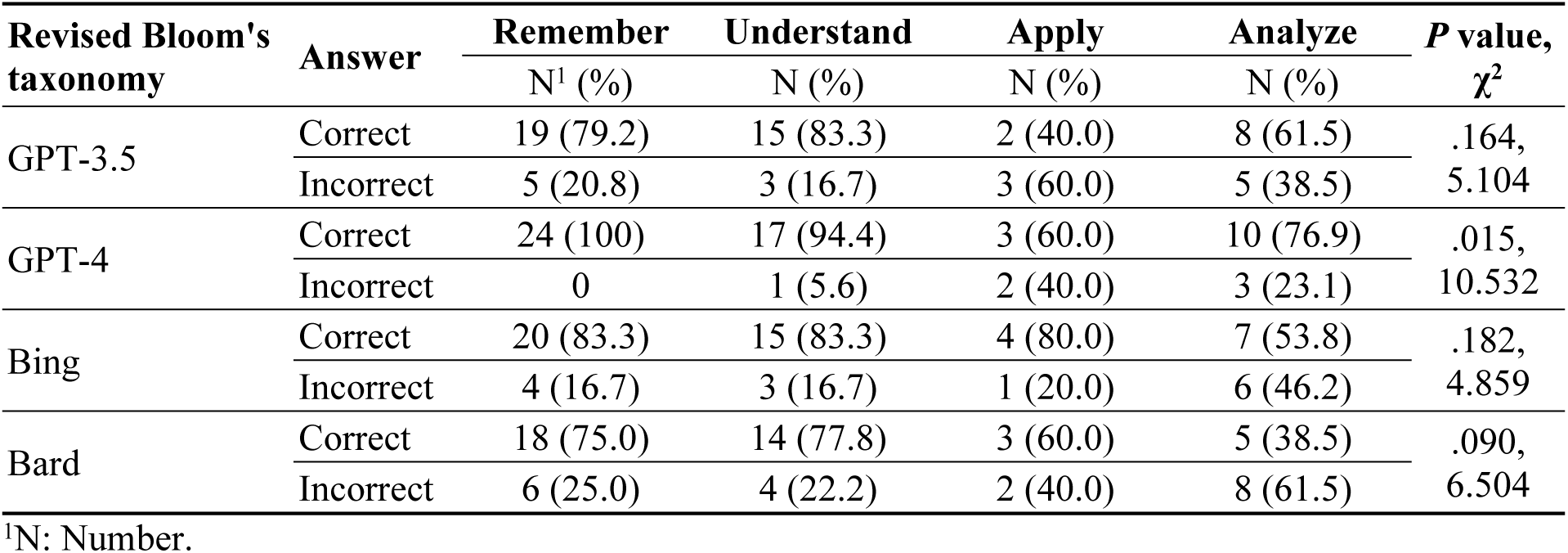
The performance of the four artificial intelligence (AI)-based models in the Clinical Chemistry multiple-choice question (MCQs) stratified per the four revised Bloom’s categories.

On the other hand, ChatGPT-3.5, ChatGPT-4, and Bard showed statistically better performance in the lower cognitive MCQs compared to the higher cognitive MCQs (**Figure 1**).

**Fig 1.**
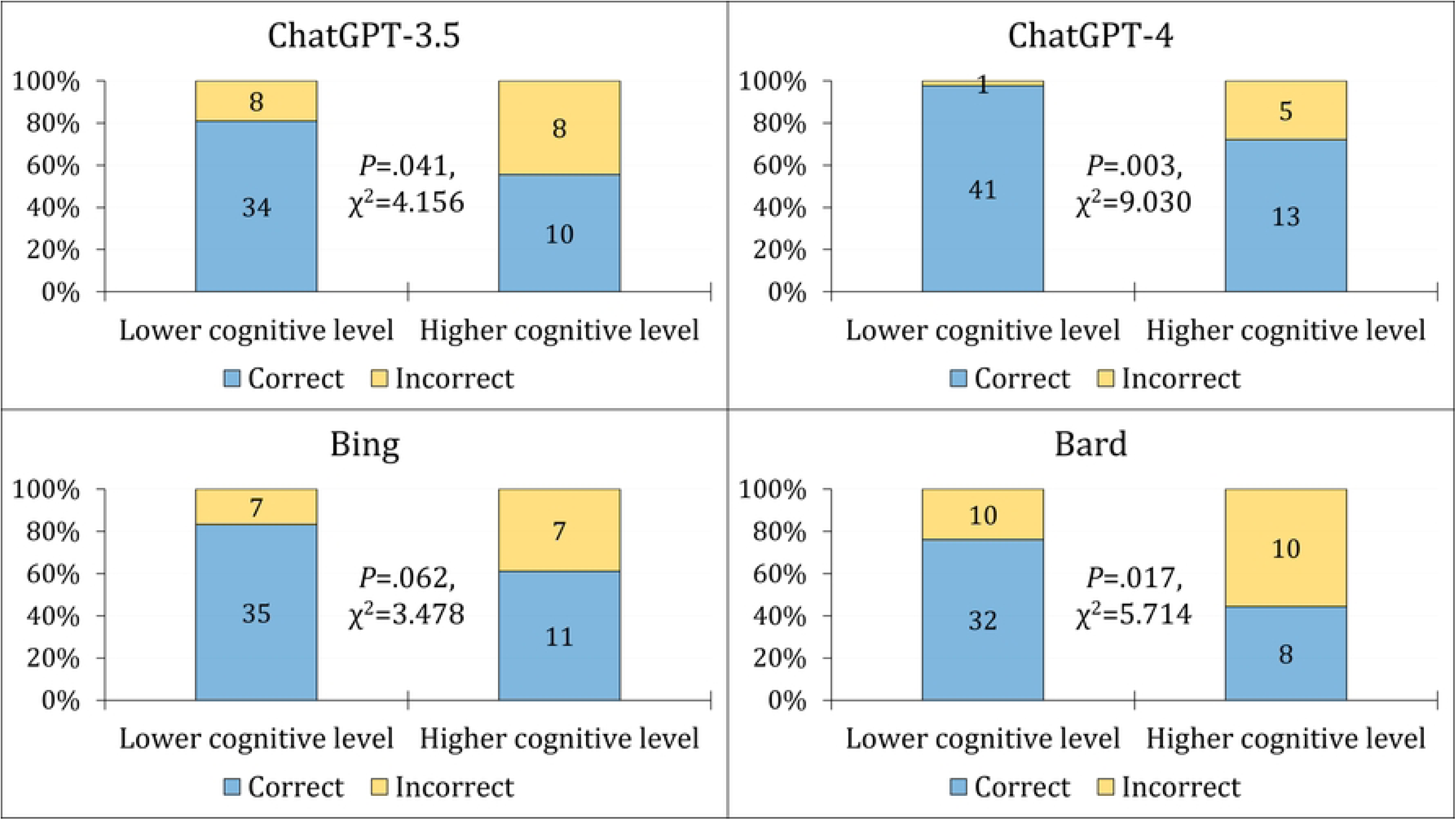
The performance of the four artificial intelligence (AI)-based models in the MCQs stratified per the revised Bloom cognitive levels.

### Performance of the AI models based on the modified CLEAR tool

In our assessment of completeness, accuracy/evidence, and appropriateness/relevance, based on the modified CLEAR tool, ChatGPT-4 was the only model rated as “Excellent” across all categories. Bing achieved an “Excellent” rating solely in appropriateness/relevance. The other AI models were categorized as “Above average” in performance (**Table 4**). The statistical analysis revealed significant superiority of ChatGPT-4 compared to the other models in all CLEAR categories, with the exception of Bing where the difference was only significant in the completeness and the overall mCLEAR score (**Table 4**).

**Table 4.** Modified CLEAR average scores for the four AI models in explaining the rationale for selecting choices.

| Assessment category | Mean±SD<sup>2</sup> | Rank | *P* value<sup>3</sup> | Post hoc test (Mann Whiteny *U* test) |  |  |  |  |  |
| --- | --- | --- | --- | --- | --- | --- | --- | --- | --- |
|  |  |  |  | GPT-3.5 vs.<br>GPT-4 | GPT-3.5 vs.<br>Bing | GPT-3.5 vs.<br>Bard | GPT-4 vs.<br>Bing | GPT-4 vs.<br>Bard | Bing vs.<br>Bard |
| ChatGPT-3.5 completeness score | 4.03±1.26 | Above average | **<.001** | **<.001** | .343 | .249 | **.001** | **.003** | .745 |
| ChatGPT-4 completeness score | 4.73±0.77 | Excellent |  |  |  |  |  |  |  |
| Bing completeness score | 4.14±1.34 | Above average |  |  |  |  |  |  |  |
| Bard completeness score | 4.19±1.18 | Above average |  |  |  |  |  |  |  |
| ChatGPT-3.5 accuracy/evidence score | 3.87±1.80 | Above average | **0.016** | **0.007** | .633 | .604 | .023 | **.002** | .324 |
| ChatGPT-4 accuracy/evidence score | 4.6±1.21 | Excellent |  |  |  |  |  |  |  |
| Bing accuracy/evidence score | 4.07±1.66 | Above average |  |  |  |  |  |  |  |
| Bard accuracy/evidence score | 3.67±1.90 | Above average |  |  |  |  |  |  |  |
| ChatGPT-3.5 appropriate/relevance score | 4.18±1.33 | Above average | 0.011 | **0.005** | .645 | .691 | .023 | **.001** | .355 |
| ChatGPT-4 appropriate/relevance score | 4.76±0.76 | Excellent |  |  |  |  |  |  |  |
| Bing appropriate/relevance score | 4.27±1.35 | Excellent |  |  |  |  |  |  |  |
| Bard appropriate/relevance score | 4.15±1.22 | Above average |  |  |  |  |  |  |  |
| ChatGPT-3.5 mCLEAR<sup>1</sup> score | 4.03±1.41 | Above average | <.001 | **<.001** | .270 | .213 | **.001** | **.002** | .868 |
| ChatGPT-4 mCLEAR score | 4.70±0.90 | Excellent |  |  |  |  |  |  |  |
| Bing mCLEAR score | 4.16±1.43 | Above average |  |  |  |  |  |  |  |
| Bard mCLEAR score | 4.00±1.41 | Above average |  |  |  |  |  |  |  |

<sup>1</sup>mCLEAR: Modified CLEAR score based on the study by Sallam et al. [38]; <sup>2</sup>SD: Standard deviation; <sup>3</sup>Calculated using the Kruskal Wallis test. Significant *P* values are highlighted in bold style.

## Discussion

The whole landscape of education, including higher education is set for a new era that can be described as a paradigm shift with the widespread and popularity of AI [13, 53, 54]. In this study, a comparison between the human and AI abilities in a highly specialized field at a high level was undertaken. Specifically, the performance of MSc students in a Clinical Chemistry exam, with an average score of 40.05±7.23 (66.75%), was used as a benchmark for comparison. Remarkably, ChatGPT-4 surpassed this human benchmark, achieving a score of 54/60 (90.00%). Bing followed with 46/60 (76.67%), outperforming both ChatGPT-3.5 (44/60, 73.33%) and Bard (40/60, 66.67%). Overall, the level of AI models’ performance underlines the advancements in AI capabilities. Additionally, these results could pave the way for a broader scientific inquiry into both the potential role of AI in educational settings as well as the usefulness of the current assessment tools in higher education.

In this study, the initial central hypothesis assumed that the human students at a postgraduate level who undertook a specialized course in a highly specialized field, namely Clinical Chemistry, would show a superior performance compared to the tested AI models. The findings of this study showed that the AI models tested not only passed the exam but showed a noteworthy performance. For example, ChatGPT-4 score equaled the highest student score and thus would be rated as an “A” student. On the other hand, the performance of the AI models in this study was not entirely an unexpected finding. This comes in light of the recent evidence showing AI models’ abilities to pass reputable exams in multiple languages such as the USMLE [37], the German State Examination in Medicine [55], the National Medical Licensing Examination in Japan [56, 57], and the Brazilian National Examination for Medical Degree Revalidation [58].

From a broader perspective, a recent systematic review highlighted the abilities of ChatGPT as an example of LLMs in various exams [35]. The review by Newton and Xiromeriti highlighted the capabilities of this popular AI model, with ChatGPT-3 outperforming human students in 11% of the included exams, with ChatGPT 4 achieving superior performance and outscoring the human performance in 35% of the included exams [35]. The current study findings were in line with the finding of better GPT-4 performance as opposed to the earlier and free GPT-3.5 version. Yet, the performance of ChatGPT-4 in comparison to the human students was noteworthy highlighting the refinements of LLMs over a short period of time.

In this study, analyzing the human students’ performance based on the revised Bloom’s taxonomy enabled elucidation of deeper insights into the assessment of cognitive aspects. The human students excelled in the “Remember” domain which is indicative of strong recalling and recognizing abilities. Additionally, the human students demonstrated a high performance in the “Understand” and “Apply” categories, with the lowest performance shown in the “Analyze” category. The lack of statistical significance in these differences suggest a balanced level of cognitive skills acquired among the students during the course despite the potential for improvement in higher-order cognitive skills entailing breakdown and organization of acquired knowledge.

On the other hand, the study findings revealed an interesting observation manifested in worse AI models’ performance across the higher cognitive domains. This observation stands in contrast to the findings of Herrmann-Werner et al., which pioneered the use of the Bloom’s taxonomy in AI model performance in MCQs [44]. Herrmann-Werner et al. demonstrated a lower level of ChatGPT performance in the lower cognitive skills in contrast to the findings of this study [44]. To the contrary, a recent study that assessed ChatGPT-3 performance in medical microbiology MCQs showed a trend similar to our findings where the AI model performed at a higher level in the lower cognitive domains [45]. This divergence of findings suggests the need for more comprehensive studies to discern the abilities of AI models in different cognitive domains, which would be helpful to guide improvements in these models and to enhance their utility in higher education.

Upon examining the performance of AI models in this study based on the MCQ metrics (FI, DE, stem and choices word count), a significant drop in performance was noted in Bing and Bard for more difficult MCQs. This finding suggests that some AI models have yet to show evolution into the level where it can handle complex queries. The absence of a correlation between MCQ stem and choice word counts and AI performance indicates that the challenge was not related to the length of the queries but rather in the inherent complexity of the prompts.

In this study, the use of the validated CLEAR tool for assessment of the quality of AI generated content presented a robust approach [38]. The rating of ChatGPT-4 as “Excellent” across all categories of completeness, accuracy/evidence, and appropriateness/relevance serves as a clear demonstration of its superiority. The Bing’s — which uses similar GPT-4 architecture — rating as “Excellent” in appropriateness/relevance was a noteworthy finding; nevertheless, the performance of this Microsoft AI model did not match ChatGPT-4 in terms of completeness and accuracy. The other AI models in this study were rated as “Above average” based on the modified CLEAR tool. This result, albeit lower than ChatGPT-4, still showed the huge potential of these freely available models, but with an evident room for improvement. The significant superiority of ChatGPT-4 over the other AI models tested in this study highlights the swift evolution of AI capabilities [59].

In the field of higher education, the implications of the study findings can be profound. The noteworthy capabilities of AI models, especially those shown by ChatGPT-4, to outperform humans at a postgraduate level could serve as a red flag necessitating the re-evaluation of traditional assessment approaches currently utilized for evaluation of students’ achievement of learning outcomes [53, 60]. Additionally, the study findings highlighted the current possible AI limitations in addressing higher-order cognitive tasks, which shows the unique value of human critical thinking and analytical skills [61]. Nevertheless, more studies are needed to confirm this finding based on a recent evidence showing the satisfactory performance of ChatGPT in tasks requiring higher-order thinking specifically in the field of medical biochemistry as shown by Ghosh and Bir [62].

Future research could focus on investigating the feasibility of integrating AI into higher education frameworks in terms of utilizing an approach that could augment the human learning (e.g., through enhancing personalized learning experience and providing instantaneous feedback) without compromising the development of critical thinking and analytical skills [5, 9, 53, 63–65]. Additionally, the ethical considerations of academic integrity should be considered in light of opportunities of academic dishonesty posed by AI models in educational settings [66–68]. This issue also extends to warrant a thorough investigation into the implications of possible decline in students’ analytical and critical thinking skills and prioritizing the human needs and value [27, 69, 70].

Finally, while the current study can provide valuable insights into the performance of AI models compared to human students in the context of Clinical Chemistry topic, several limitations should be considered when interpreting the results. Future research in this area would benefit from addressing these limitations that included: First, this study employed a limited dataset of 60 MCQs. This limited number of MCQs inherently restricts the scope of performance evaluation. Second, the use of the CLEAR tool, albeit standardized, introduces a subjective element in evaluating the content generated by AI models. This subjectivity could lead to a potential bias in the assessment of AI responses if approached by different raters.

Thus, the AI content evaluation was not entirely devoid of subjective judgment despite the use of key answers to reduce this subjectivity bias. Third, the exclusive concentration on Clinical Chemistry as a subject is both a strength and a limitation. While it allowed for a deep insight this specific health discipline, it limits the generalizability of the findings to other academic fields, since different subjects may present unique challenges that were not addressed in this study. Fourth, LLMs are evolving rapidly, and this study only provided a snapshot of AI models’ performance at a specific time point. Therefore, this study may not fully represent the potential improvements or advancements in AI capabilities that have occurred or may occur shortly after the study period. Fifth, the exam metrics, derived from the performance of a limited number of students (*n*=20), might have been influenced by various external factors. These include the format of the exam and its time limits and the specific cohort of students.

In conclusion, the current study provided a comparative analysis of the human versus AI performance in a highly specialized academic context at the postgraduate level. The results could motivate future research to address the possible role of AI in higher education reaping its benefits while avoiding its limitations. The ideal approach would be to use the strengths of AI as a complement to the unique capabilities of human intellect. This can ensure the evolution of the educational process in an innovative way aiding in students’ intellectual development. Importantly, the study results call for a revision of the current assessment tools in higher education with a focus on improving the assessment of higher cognitive skills.

## Acknowledgments

NA.

## Data availability statement

The data that support the findings of this study are available on request from the corresponding author (M.S.). The data are not publicly available due to the confidentiality of the questions created for an exam purposes.

## Author contribution

**Conceptualization:** Malik Sallam

**Data Curation:** Malik Sallam, Khaled Al-Salahat

**Formal Analysis:** Malik Sallam

**Investigation:** Malik Sallam, Khaled Al-Salahat, Huda Eid, Jan Egger, Behrus Puladi

**Methodology:** Malik Sallam, Khaled Al-Salahat, Huda Eid, Jan Egger, Behrus Puladi

**Project administration:** Malik Sallam

**Supervision:** Malik Sallam

**Visualization:** Malik Sallam

**Writing – Original Draft Preparation:** Malik Sallam

**Writing – Review & Editing:** Malik Sallam, Khaled Al-Salahat, Huda Eid, Jan Egger, Behrus Puladi

